# Living *hi*: GBMSM narratives of pleasure, risk, and everyday *hi-fun* (chemsex) navigation in Thailand

**DOI:** 10.64898/2026.03.19.26348853

**Authors:** Worawalan Waratworawan, Nattharat Samoh, Alison J Rodger, Harry Prabowo, Gloria Lai, Nittaya Phanuphak, Siripong Srichau, Verapun Ngamee, Adam Bourne, Thomas E. Guadamuz, T Charles Witzel

**Affiliations:** Mahidol Center for Health, Behavior and Society, Faculty of Tropical Medicine, Mahidol University, Bangkok, Thailand; Institute for Global Health, University College London, London, UK; Asia Pacific Network of People Living with HIV/AIDS, Bangkok, Thailand; International Drug Policy Consortium, Bangkok, Thailand; Institute for HIV Research and Innovation, Bangkok, Thailand; APCOM Foundation, Bangkok, Thailand; Ozone Foundation, Bangkok, Thailand; Australian Research Centre in Sex, Health and Society, La Trobe University Melbourne, Victoria, Australia

**Keywords:** Gay, bisexual and other men who have sex with men (GBMSM), Hi-Fun, Sexualised drug use, Thailand

## Abstract

**Background:** *Hi-fun* among gay, bisexual and other men who have sex with men (GBMSM) is increasing across Southeast Asia. Yet research remains focused on epidemiological trends and health risks, with limited qualitative work capturing lived experiences, motivations and support needs. These gaps are pronounced in Thailand, where social, legal and cultural structures emphasise control and enforced abstinence over harm reduction and holistic support.

**Methods:** This qualitative study drew on five FGDs and six IDIs with 30 Thai and non-Thai GBMSM in Bangkok, Khon Kaen and Pattaya. Data explored contexts, experiences, strengths, challenges and support needs related to *hi-fun*. Thematic analysis was conducted, and findings were reviewed with a community advisory board and partner NGOs.

**Results:** Participants described *hi-fun* as a distinct world enabling self-expression, pleasure, intimacy and a sense of being valued and accepted beyond what everyday life affords. This world was shaped through equipment preparation, participant screening and unwritten rules promoting trust and safety. *Hi-fun* offered emotional and social benefits, including connection and belonging. However, participants also reported risks, including mental health challenges, dependence, sexual performance difficulties, violence, non-consensual recording, financial strain and legal consequences. *Hi-fun* became problematic when individuals lost control, encountered police or mandatory rehabilitation, experienced health decline or struggled with daily functioning. Participants emphasised needs for sterile equipment, post-use care, accurate information, stigma-free environments and legal reforms aligned with lived realities.

**Conclusion:** Supporting GBMSM engaged in *hi-fun* in Thailand requires moving beyond abstinence-focused approaches toward flexible harm reduction and structural reforms that uphold dignity, safety and equity.

## Background

The phenomenon of *hi-fun* among GBMSM in Thailand, often called chemsex in Western countries, has gained increasing attention in academic, health system and media discussions. (Guadamuz & Boonmongkon, 2018; Nevendorff et al., 2021; Thomson Reuters Foundation, 2021) *Hi-fun* refers to “the intentional combining of sex and crystal methamphetamine (ice) to enhance intimacy and pleasure with one or more other man/men, facilitated by technology and usually in a private setting.” (Witzel et al., 2025). These practices often occur in informal but prearranged contexts organised through dating apps and may involve multi day sex parties. Many participants intentionally use substances to increase confidence, disinhibition, and openness to more intense sexual experiences. (Tan et al., 2018).

Existing research has helped identify several health implications of *hi-fun* including heightened risks of HIV and sexually transmitted infections (STIs) acquisition, sexual violence, mental health concerns, and the need for health-promoting activities for substance-using GBMSM (Cheung et al., 2024; Hibbert et al., 2019; Strong et al., 2022; Witzel et al., 2023). Indeed, much of the current literature relies on risk based framings and pays limited attention to the social, emotional, and the cultural meaning attached to *hi-fun* (APCOM, 2021; Møller & Hakim, 2023; Wang et al., 2023). Although some studies in high-income countries have begun to explore pleasure, agency, and lived experience in sexualized drug use (Bryant et al., 2018; Møller & Hakim, 2023; Nagington, 2024), such research remains limited in low-and middle-income settings, especially in Southeast Asia where the voices of people with lived experience remain limited. This constrains the development of interventions which acknowledge the pleasure and value derived from in *hi-fun*, factors shown to support more engaging and effective service provision. (Hawkinson et al., 2024)

Exploring *hi-fun* in Thailand is further complicated by sensitive of drug use, sexuality, and gender sexual diversity in public discourse. When publicly discussed, they are often accompanied by moralistic framings and stigma, reinforced by criminalization, compulsory treatment systems, and moral narratives that position drug use as deviance rather than a health issue. (Vatanasin & Jinjutha Chaisena Dallas, 2021; Yangyuen et al., 2020).

In Thailand, family and social expectations rooted in patriarchal and patrilineal values add further pressure. In Thai Chinese families, sons are regarded as responsible for maintaining male lineage, while daughters marry out of the family (Jackson, 2011; Pongtriang et al., 2017). These expectations, strengthened by notions of filial piety, place heavy burdens on sons to meet familial and social obligations (Bao, 2004; Guadamuz & Boonmongkon, 2018; Kongjareon et al., 2022; Ojanen et al., 2020; Shrestha et al., 2020). This results in many GBMSM living under often intense pressure from families, health systems, and state policies that leave little room for personal desires, pleasure, or self-defined ways of managing life.

In policy settings, *hi-fun* is often reduced to a biomedical health issue, overshadowing social, cultural, and community contexts, as well as the benefits individuals describe in their engagement with sexualised drug use (EpiC & APCOM, 2021). Although the United Nations Office on Drugs and Crime has urged member states to shift from punitive drug approaches toward models grounded in human rights (UNODC, n.d.), the Thai legal system has yet to fully realize such reforms. Persistent fragmented and inconsistent legislation creates barriers for marginalized group. For example, repeated revisions methamphetamine possession thresholds have created uncertainty in both interpretation and enforcement. Lower thresholds increase the likelihood that individuals possessing drugs for personal use are misclassified as traffickers, since legal limits do not reflect real world consumption patterns (Lai, 2024). This ambiguity heightens legal risk for *hi-fun* participants and discourages them from seeking healthcare or support. Methamphetamine and similar substances remain heavily criminalized, further contributing to reluctance in accessing services (Witzel et al., 2023).

Biomedical paradigms shaping current knowledge surrounding *hi-fun* tend to emphasize treatment or harm reduction by positioning users as patients in need of management rather than respecting them as people with narratives, desires, and their own strategies for living. Viewed through the lens of Critical Medical Anthropology (Farmer, 2010), these structures often silence vulnerable groups through forms of structural violence including social exclusion, stigma, and unequal access to resources (Scheper-Hughes, 2004). By framing *hi-fun* strictly in terms of disease and risk, complexity is reduced, and dominant systems reinforce definitions of abnormality. Although harm reduction has gained recognition, much discourse remains governed by medicalised thinking that separates drug use from social and cultural realities. Without listening to people with lived experience, support systems risk reinforcing power imbalances and failing to uphold dignity and human need (Farmer, 2010).

This study seeks to create meaningful space for the authentic voices of people with *hi-fun* experience. It explores how individuals define sexualised drug use, interpret their pleasures, risks, and vulnerabilities, and describes the kinds of support that best align with their lived contexts. We draw on the diverse settings of Bangkok, Khon Kaen, and Pattaya, recognising that no single narrative captures the experiences of all participants and avoiding reductions into external or dominant frameworks (Foucault, 1978). By grounding analysis in lived realities, the research outlines supportive approaches including harm reduction strategies that uphold physical safety along with dignity, agency, and human needs.

This qualitative study therefore aims to explore the experiences and perspectives of GBMSM who engage in *hi-fun* and other types of sexualised drug use, including their strengths and challenges. It seeks to identify the types of support they desire in order to contribute to the development of harm reduction strategies grounded in the everyday contexts of GBMSM communities in Thailand.

## Methods

We conducted a qualitative study using focus group discussions to explore community norms, supplemented by in-depth interviews to add depth and capture perspectives of GBMSM with privacy concerns.

### Coproduction

The study was co-produced with a community advisory board comprising representatives from policy, clinical, and community organisations, as well as regional networks of people living with HIV and people who use drugs. This group contributed to developing instruments, recruitment strategies, and the analysis and interpretation of findings.

### Selection and Recruitment

Participants were recruited through NGOs experienced with GBMSM in three Thai locations: Bangkok with the Institute of HIV Research and Innovation, Khon Kaen with ACTTEAM and Pattaya with the Health and Opportunity Network, and through investigators’ social networks. Eligible participants were cisgender or transgender men aged at least 18 who reported using stimulant or psychoactive substances such as crystal methamphetamine, GHB or GBL, ketamine, MDMA, cocaine or other stimulants before or during sex with another man within the past 12 months. Although no sampling frame was used, we required that at least 20% of the sample be non-Thai born to reflect the diversity of GBMSM communities in Thailand.

The three study sites were selected for their distinct social and cultural contexts. Bangkok represents an urban centre attracting Thai and international GBMSM. Khon Kaen reflects a regional multicultural setting shaped by students, workers and migrant labourers. Pattaya is a tourism-driven city linked to the service industry and transactional sex. These varied environments shape different GBMSM experiences.

### Data Collection

Topic guides for focus groups and interviews addressed issues related to sexualised drug use including hi-fun, motivations, challenges and support needs (Witzel et al., 2025). Participants received information about study aims, procedures, confidentiality and withdrawal right several days before data collection through NGO staff. Additional clarification was provided as needed.

Verbal informed consent was obtained and audio recorded. Researchers explained the study purpose, risks and data protection measures before beginning each session. To minimise stigma and discomfort, anonymity was prioritised and no names were collected unless required for recruitment; these were destroyed promptly. A short anonymous demographic survey captured age, gender identity, education, HIV status, PrEP use, substances used during sex and country of birth.

During data generation participants were referred to by numerical codes and reminded not to use names when discussing experiences. They were invited to reflect on strengths and challenges of *hi-fun* and identify priority issues. Data collection was conducted by three trained researchers with expertise in ethics and qualitative interviewing (WW, NS & TCW). Sessions were held at NGO/University offices. Participants received 1,000 THB compensation.

### Data Analysis

Audio files were transcribed verbatim, translated where necessary, reviewed and anonymised. Thematic analysis followed established procedures (Terry et al., 2017). WW and TCW reviewed the transcripts to develop preliminary themes, which were then tested and refined during coding. New themes were incorporated when researchers reached consensus. Two researchers independently coded all transcripts using NVivo, and all data were managed within the UCL Data Safe Haven. The research team subsequently consolidated the key themes and shared preliminary findings with the advisory board and NGO partners for feedback and triangulation.

### Ethics Approvals

Ethical approval was granted by the Mahidol University Research Ethics Committee (COA 2024/030.1902) and the University College London Research Ethics Committee (COA 24583/001).

## Results

We conducted five FGDs and six IDIs with 30 participants across Bangkok (n=12), Khon Kaen (n=9), and Pattaya (n=9). Five focus groups were conducted with 2–9 participants each. Most identified as gay (83%, n=25) and had completed at least a bachelor’s degree (60%, n=18). Nearly all were aged 26–45 (93%, n=28). One third were living with HIV; among HIV-negative participants, nine were using PrEP. Crystal methamphetamine was the most used substance in the past 12 months (n=23), followed by MDMA (n=14), ketamine (n=12), cocaine (n=10) and GHB/GBL (n=5). Most were Thai nationals (77%, n=23); seven were born abroad (23%). See Table 1 for full demographics.

**Table 1:** Participant demographics.

| **Variables** | **N=30** |
| --- | --- |
| **Age** |  |
| 18-25 | 1 |
| 26-35 | 16 |
| 36-45 | 12 |
| 46+ | 1 |
| **Sexual orientation** |  |
| Gay | 25 |
| Bisexual | 5 |
| **Born in Thailand** | 23 |
| **Born in other country** | 7 |
| **HIV status** |  |
| Negative | 6 |
| Negative/taking PrEP | 9 |
| Living with HIV | 10 |
| Prefer not to say | 5 |
| **Education qualification** |  |
| Bachelor's degree or higher | 18 |
| Secondary school | 3 |
| Below secondary school | 9 |
| **Location** |  |
| Bangkok | 12 |
| Khon Kaen | 9 |
| Pattaya | 9 |
| **Drugs taken in prior 12 months** |  |
| Cocaine | 10 |
| Ecstasy/MDMA | 14 |
| GHB/GBL | 5 |
| Ketamine | 12 |
| Crystal methamphetamine | 23 |

Findings are presented in six themes: (1) setting the scene and exploring the context of *hi-fun* environments; (2) understanding pleasure, intimacy, and identity as core strengths; (3) behind the *hi*: experiences, concerns and risks participants encounter; (4) when *hi-fun* becomes a problem; (5) managing substance use and strategies for minimizing harm; and (6) identifying support needs and desired approaches beyond abstinence-based interventions.

### Setting the scene: Inside the world of *hi-fun*

Over half the participants described *hi-fun* as entering a distinct world—a space separate from everyday life where they could express themselves freely and experience joy, authenticity and escape. Rather than impulsive or chaotic, this world was intentionally created through careful preparation. Participants described obtaining substances, especially *ice* (crystal methamphetamine), along with tools such as syringes, saline solution and sexual-enhancement products. In Bangkok, access to items like saline was limited, sometimes leading to higher resale prices, particularly at night.

Sessions often lasted two to three days, typically on weekends or holidays. Participants stocked snacks and sugary drinks and preferred not to leave the space once inside, reinforcing the sense of a contained *hi-fun* world designed for continuity and immersion.

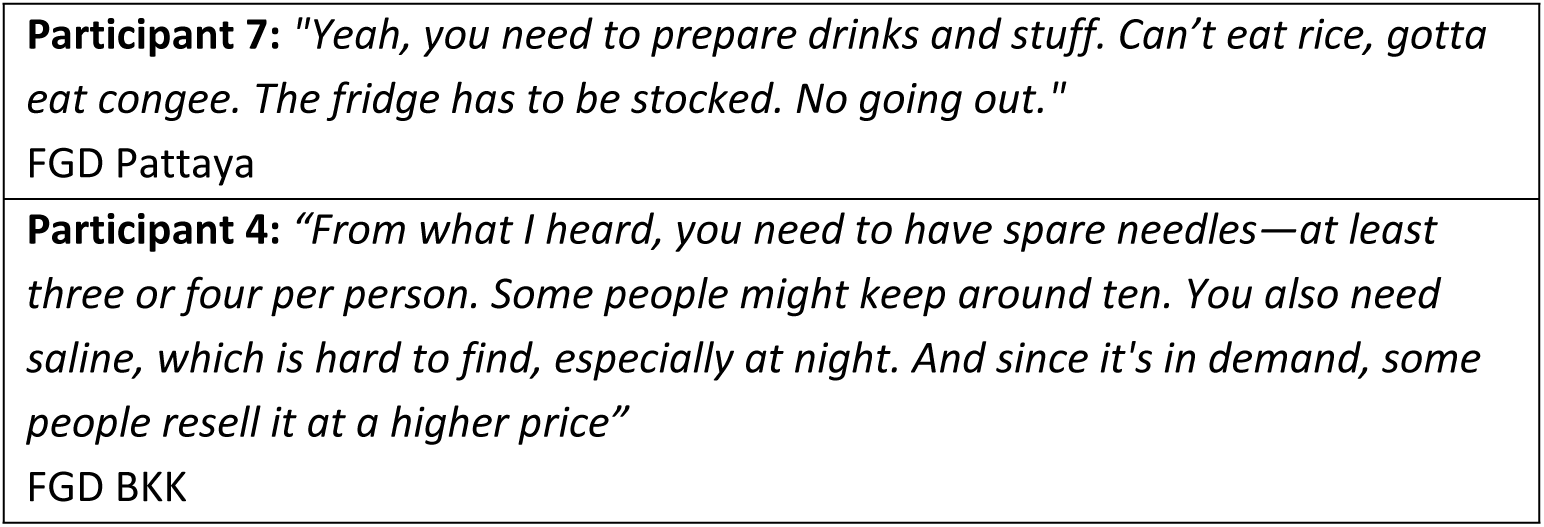

Selecting the right people was central to shaping this world. Participants screened potential partners to manage risks such as undercover policing, theft or incompatible dynamics. Screening through photos or video of the face, body or genitals allowed participants to assess attraction, compatibility, perceived safety and mutual sexual interest. These appearance-based evaluations frequently determined who was considered suitable for joining a session. When compatibility was strong, the world of *hi-fun* felt pleasurable and emotionally engaging; when it was not, participants often withdrew to avoid disrupting the experience.

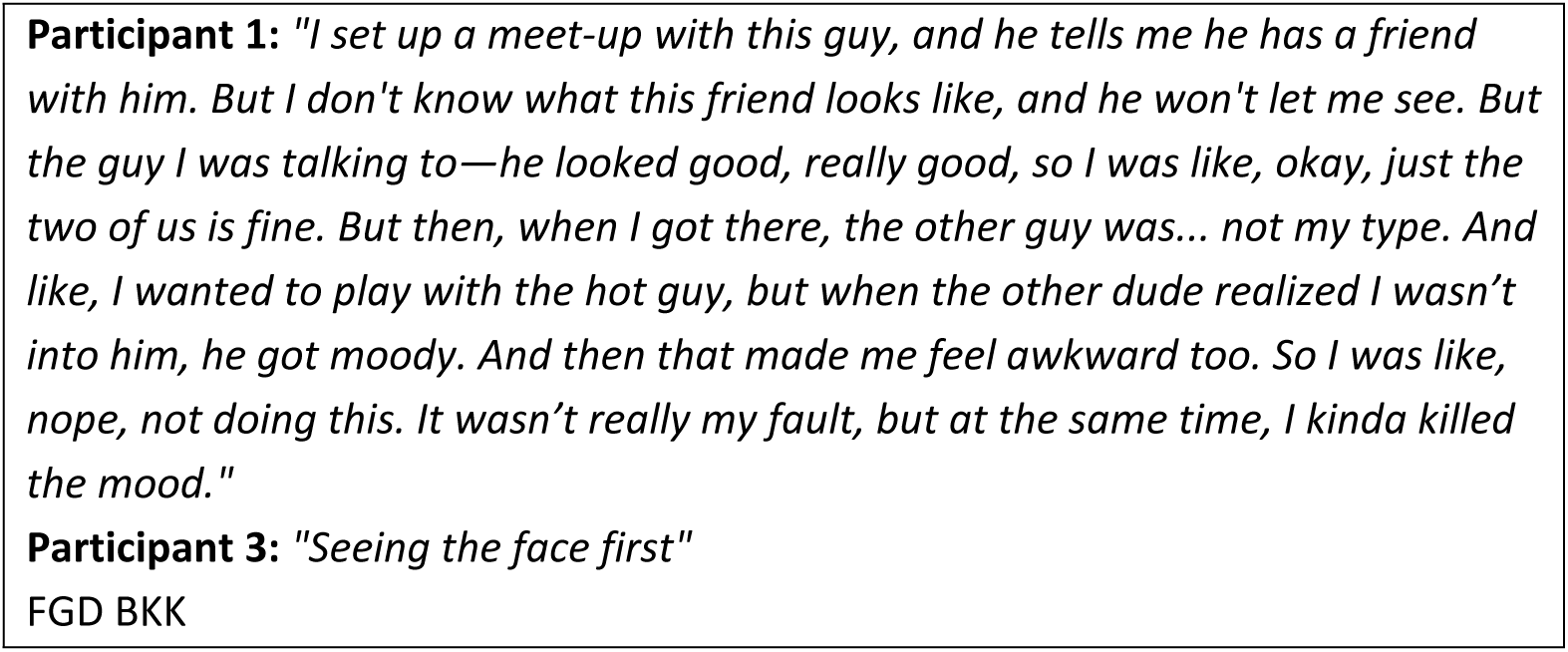

Atmosphere was another deliberate part of constructing the *hi-fun* world. Participants shaped lighting, sound and environment to cultivate mood, intimacy and safety. Soft or dim lighting was preferred for privacy and confidence. Some used music or television to sustain arousal, while others preferred silence to avoid attention, especially in hotels. In Khon Kaen, participants often used resorts or hotels because hosting at home risked exposure in tight-knit communities; thus, managing the sensory environment was essential to sustaining the protected world they created.

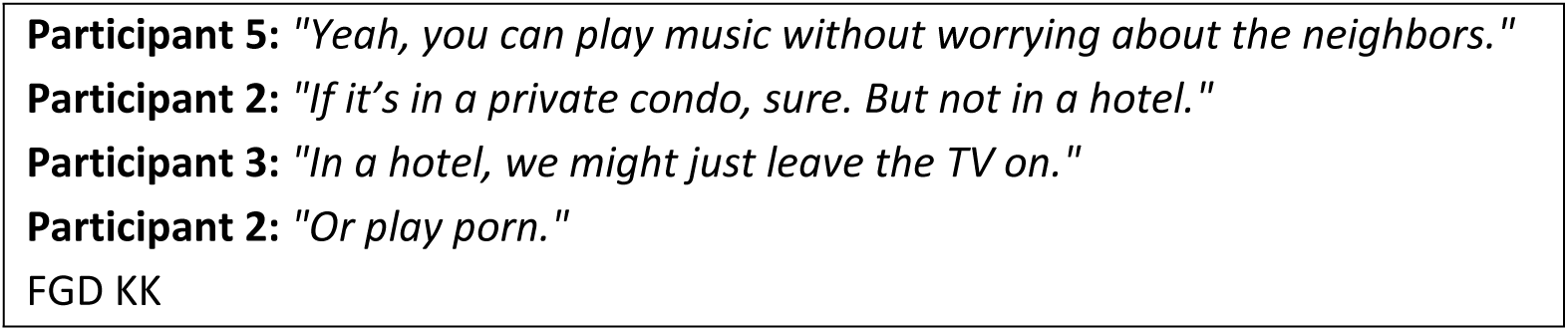

Participants also described unwritten normative rules that governed behaviour while maintaining trust and cohesion. Leaving early could raise suspicion, and phone use was discouraged because it disrupted emotional connection and could be perceived as a security threat. Shared expectations around substance use also reinforced collective participation: many emphasised that *hi-fun* was a shared experience and that using the same substances in the same ways contributed to the sense of being in a unified, mutually dependent world.

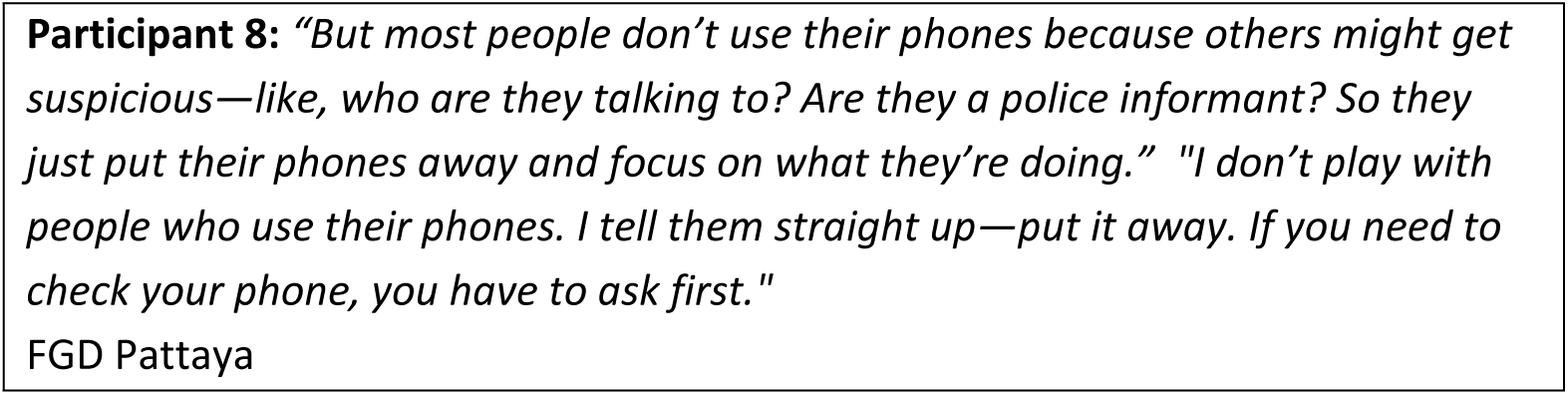

### Pleasure, community, and identity: *Hi-fun* strengths

Many participants described *hi-fun* as offering more than sexual pleasure. It functioned as an emotional and psychological space of release where they could reduce stress, ease loneliness and express themselves without judgment, which was particularly important in social contexts where sexual minorities often feel marginalised. Using ice heightened excitement, prolonged sexual pleasure and created experiences that felt distinct from everyday encounters. In Bangkok, participants highlighted the intensity of sex and how substances deepened emotional connection. In Pattaya, *hi-fun* was also seen as a way to cope with work stress and pressures tied to the tourism and sex-work economy. Across sites, *hi-fun* emerged as a world where participants could restore themselves, explore identity and access forms of empowerment unavailable in their daily lives. Participants also described feeling desired, chosen and affirmed within *hi-fun* sessions, in ways rarely available in their everyday lives.

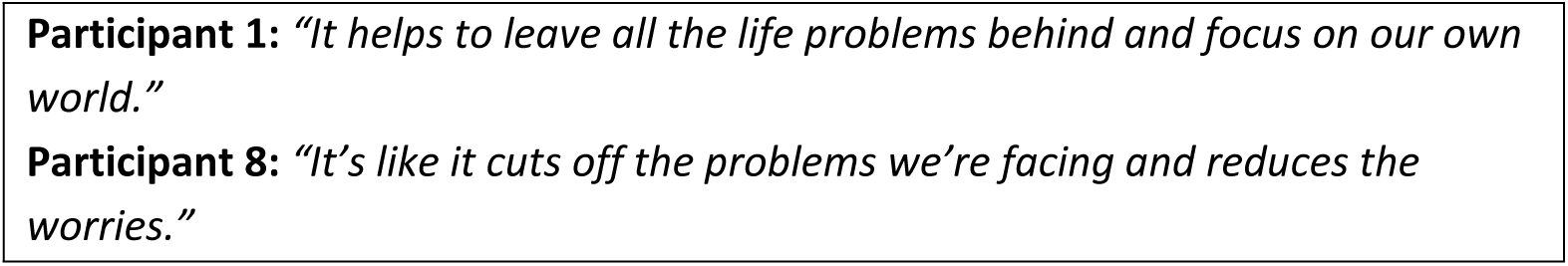
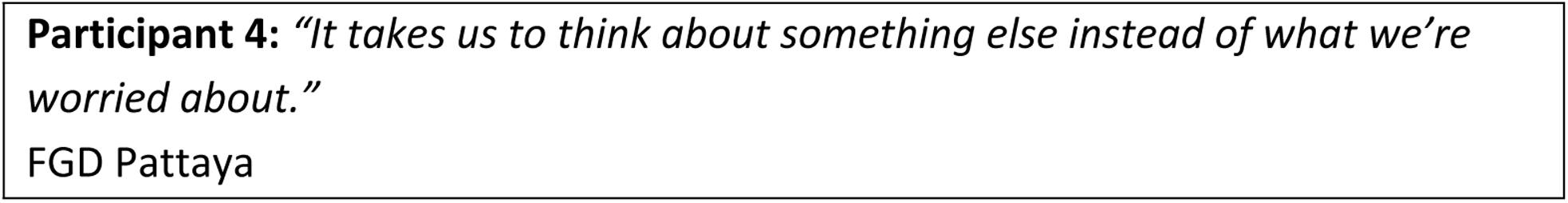

Some participants described gaining skills through *hi-fun* participation. These included safer injecting practices, caring for friends in cases of overdose, and learning how to manage drug interactions and bodily responses. Regular injectors developed expertise in preparing equipment, choosing injection sites and administering basic first aid. Such knowledge reflects a form of community resilience built within *hi-fun* spaces.

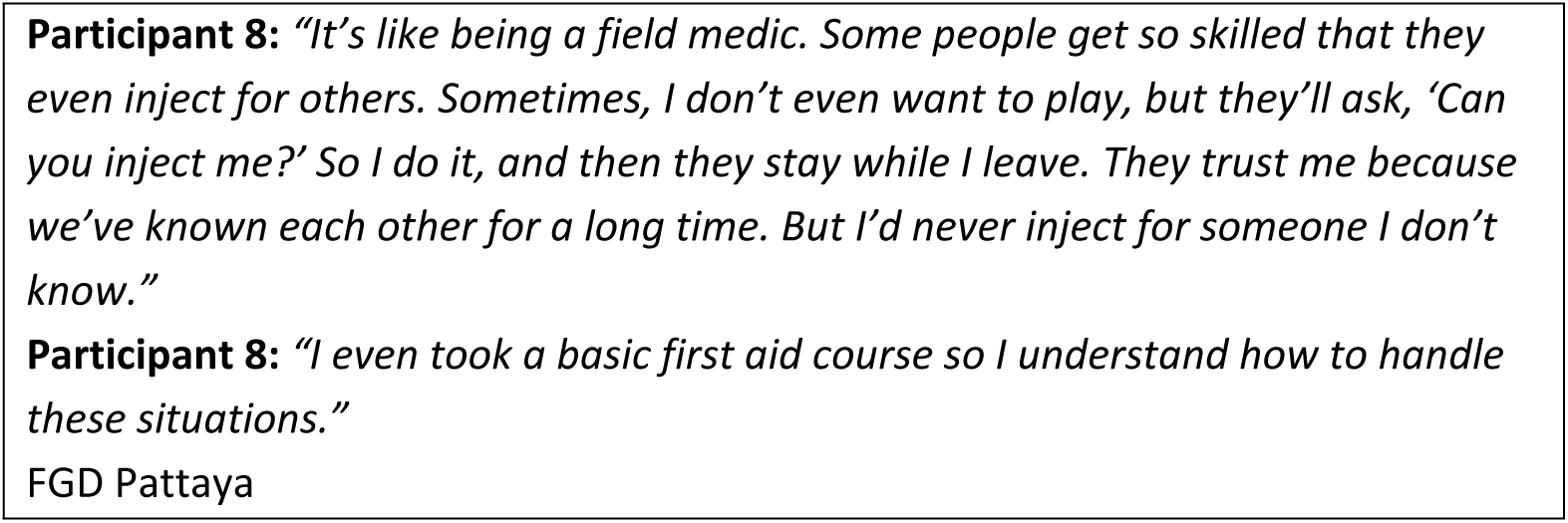

Despite these skills, mutual care was not consistently present. A smaller number of participants recalled receiving support during early drug use or emergencies. In Khon Kaen, *hi-fun* was described as intimate and personally meaningful, but opportunities for group care were limited. These accounts reveal that while *hi-fun* can foster self-reliance and occasional networks of care, access to such support varies considerably across groups.

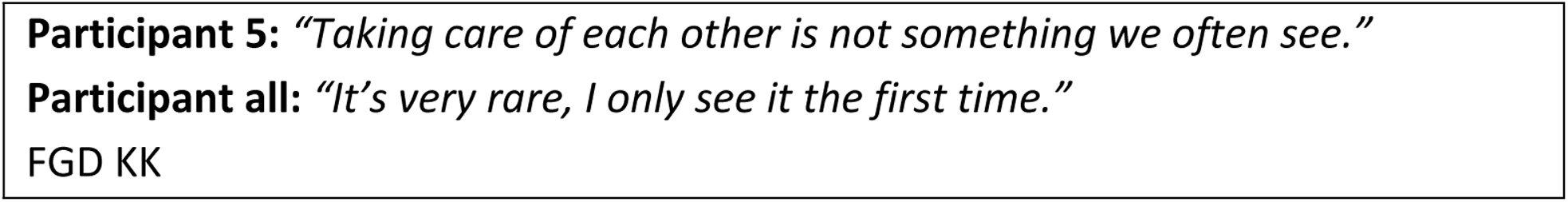

### Behind the *hi*: experiences, concerns and risks participants encounter

Many participants described mental health challenges as one of the most significant but least discussed difficulties linked to *hi-fun*. Experiences such as loss of control, paranoia, hallucinations, impaired judgment and emotional withdrawal were common, alongside intense comedowns that sometimes triggered deep distress. In Pattaya, these emotional crashes were often intertwined with romantic or relationship difficulties and, for some, led to thoughts of self-harm.

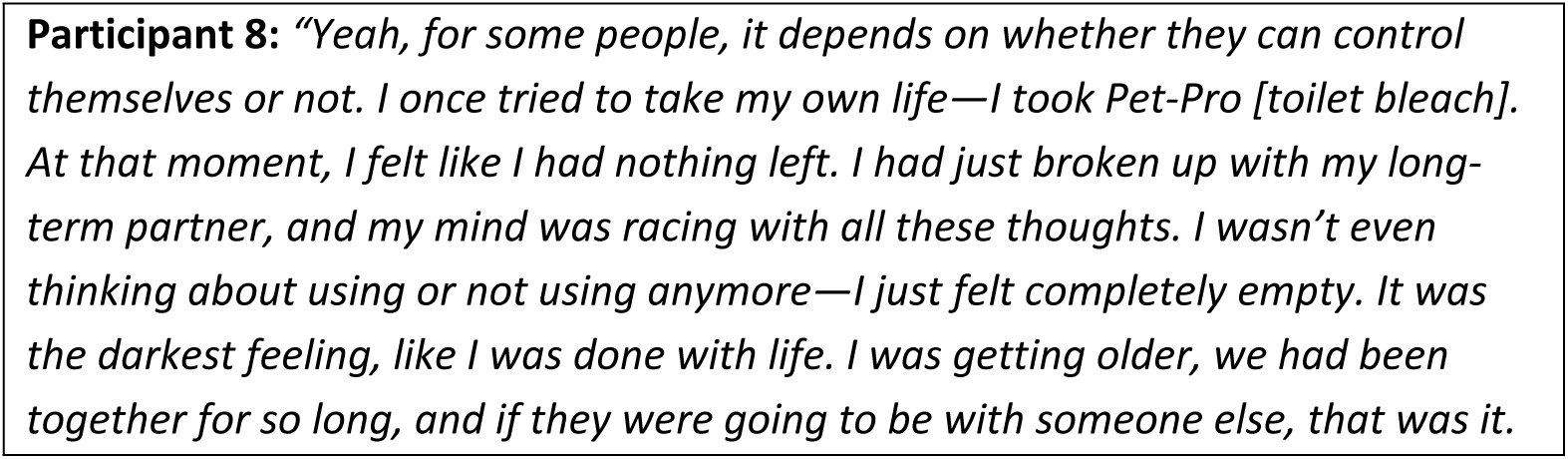
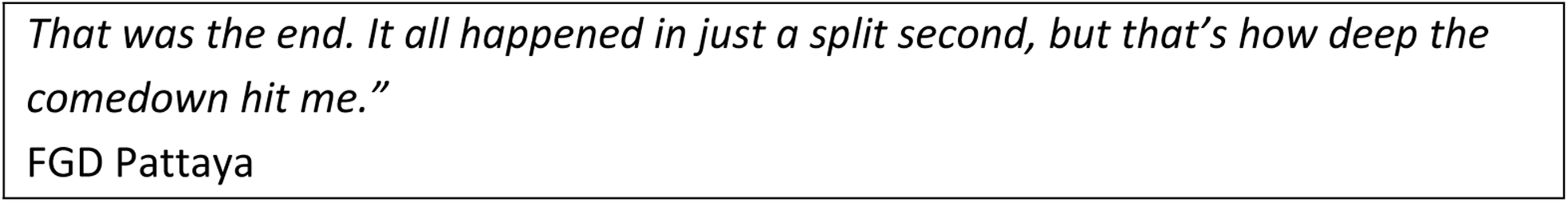

Additionally, some participants also expressed concerns that sex without substances no longer felt satisfying after experiencing the heightened pleasure, confidence and intimacy associated with *hi-fun*. Some felt their sexual identity and fulfilment became dependent on drug-enhanced encounters.

Erectile dysfunction was another prominent concern, especially among frequent users. Because sex was central to *hi-fun*, difficulties maintaining an erection could disrupt the session and generate anxiety, leading many to rely on medications like Viagra to sustain performance. These experiences shaped feelings of confidence, self-image and the overall quality of hi-fun encounters.

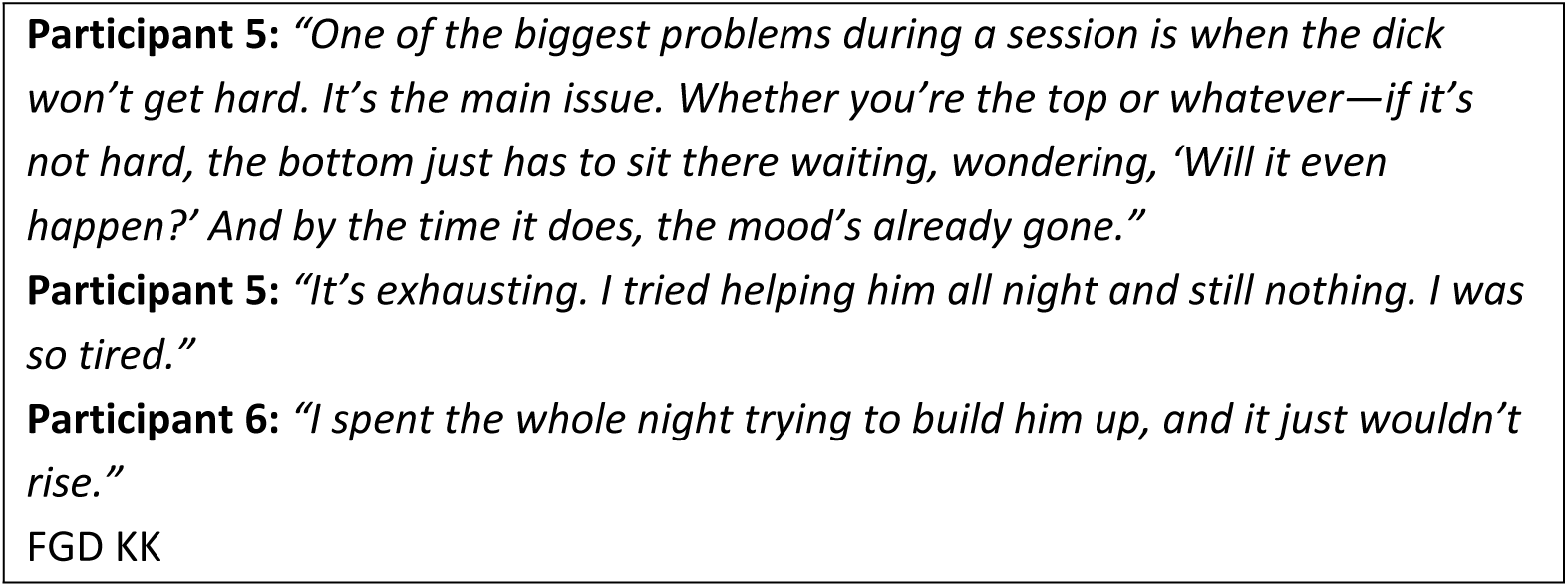

Participants also worried about physical and sexual health risks, including potential exposure to STIs, fatigue, insomnia and weight loss. Some consequences appeared immediately after use; others emerged with prolonged engagement. While less frequently discussed, overdose, cardiac issues and injecting-related harms were acknowledged as concerns.

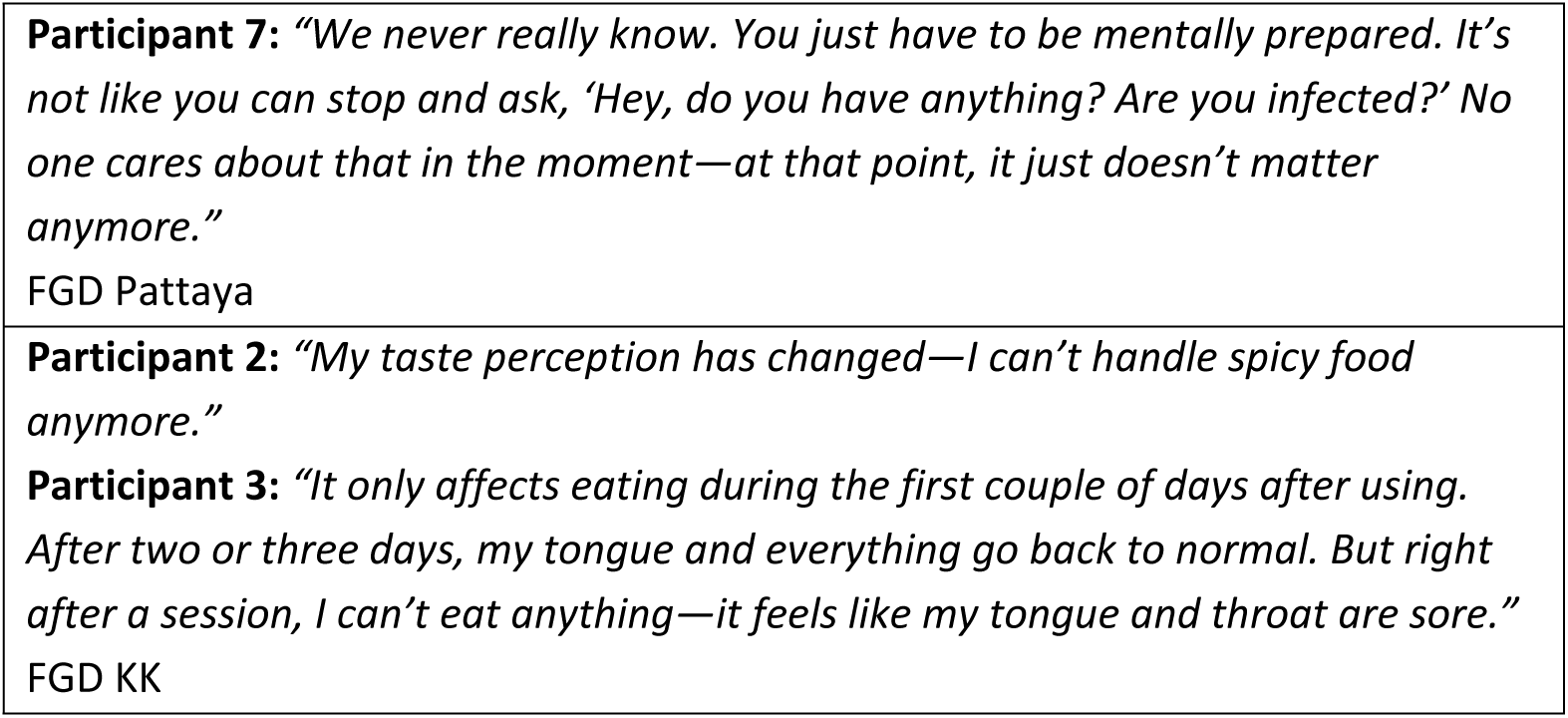

Legal risks added another layer of fear. Because drug use in Thailand carries criminal penalties and can result in mandatory rehabilitation, many participants experienced ongoing paranoia about police inspections, raids or undercover officers. Some described being searched, arrested or pressured to pay bribes. These fears made *hi-fun* feel unpredictable and emotionally taxing.

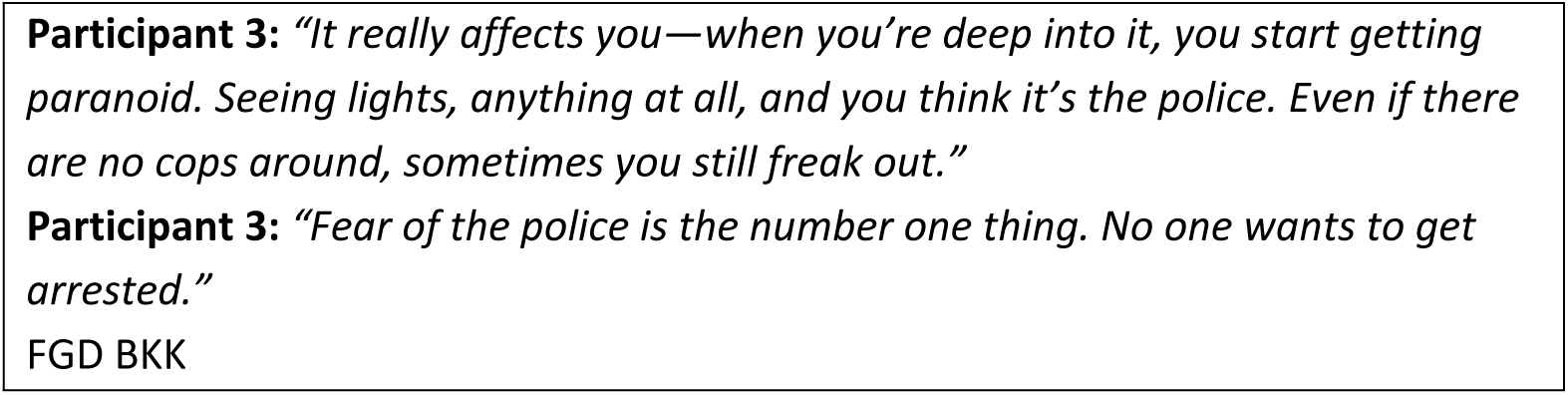

Safety concerns extended beyond law enforcement. Participants described risks of violence, theft, deception and non-consensual recording, particularly when meeting unfamiliar partners. The unpredictability of others’ behaviour contributed to a sense of vulnerability, and some men prepared strategies in advance to manage conflicts or unexpected incidents.

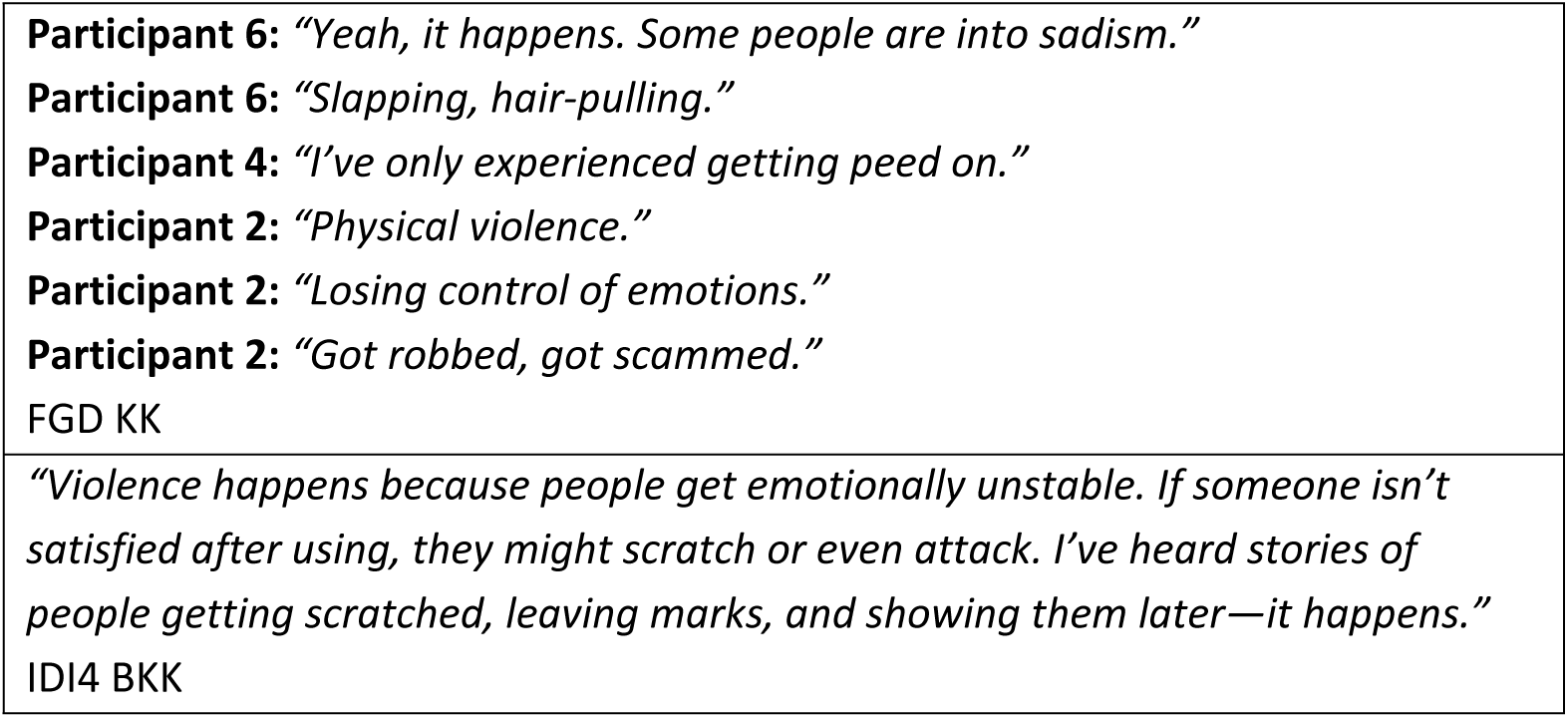

Finally, participants noted the financial strain associated with *hi-fun*, including costs for drugs, accommodation, equipment and transport. Some turned to additional income sources, gambling or pawning belongings in order to continue participating.

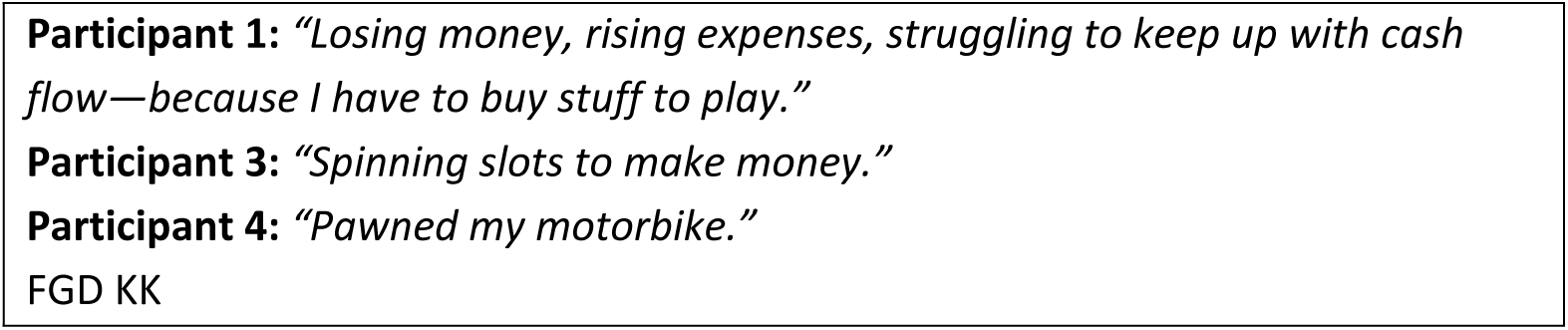

### Beyond control: When *hi-fun* becomes a problem

In addition to the risks linked to *hi-fun*, participants described several situations in which *hi-fun* shifted from pleasurable to problematic. A common indicator was an increase in frequency beyond one’s personal limits. For many, engaging more often than intended signaled a loss of control and raised concerns about dependence.

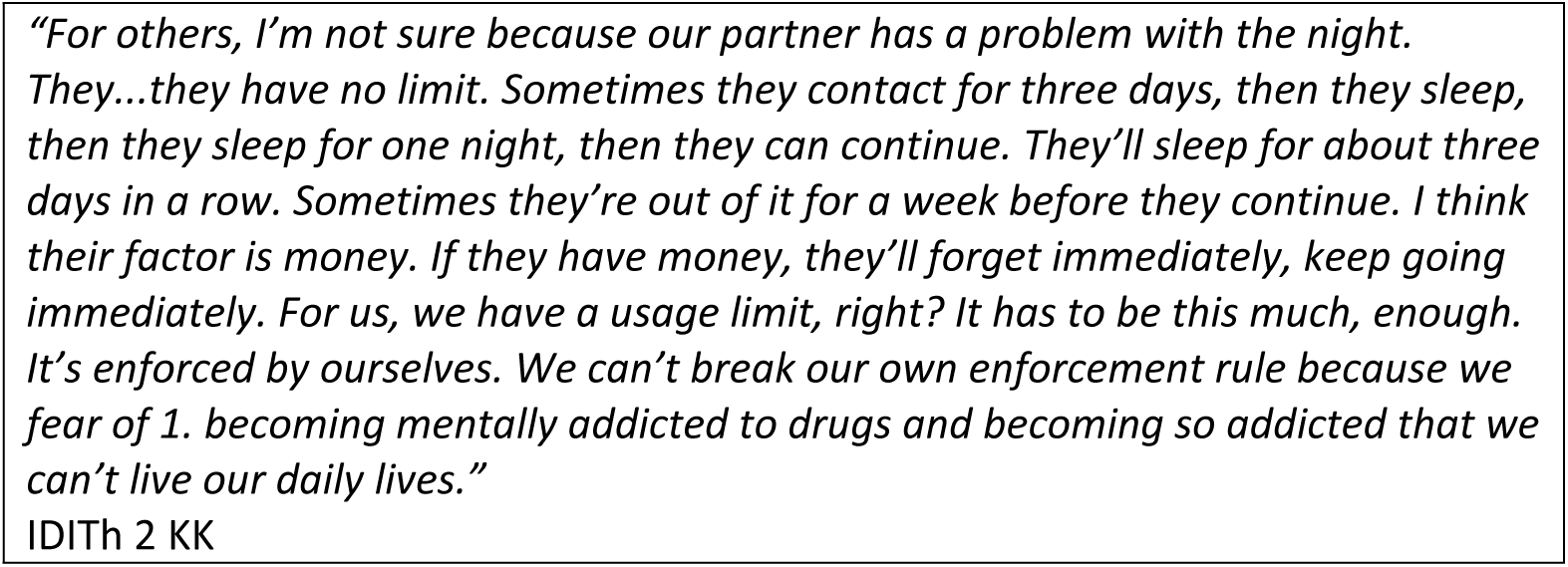

Loss of control extended beyond frequency. In Bangkok, participants described difficulties regulating their behavior during sessions, including distorted perceptions, risky interactions or actions they would not normally take. When these behaviors emerged, men began to view their *hi-fun* use as problematic.

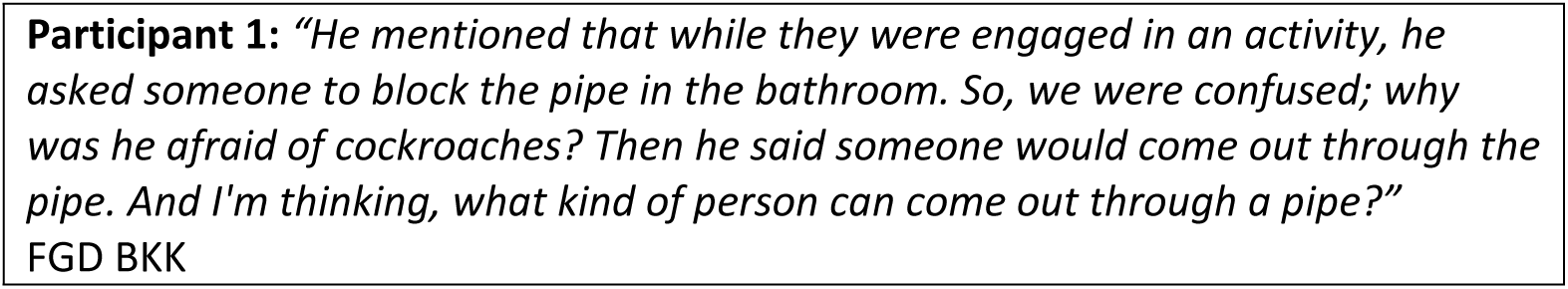

Being unable to recall events during *hi-fun* was another sign of concern. Participants also noted that social withdrawal, reduced interaction with others and declining work performance indicated that *hi-fun* was negatively affecting their daily lives. In Khon Kaen, men often linked these consequences to financial strain and deteriorating physical health.

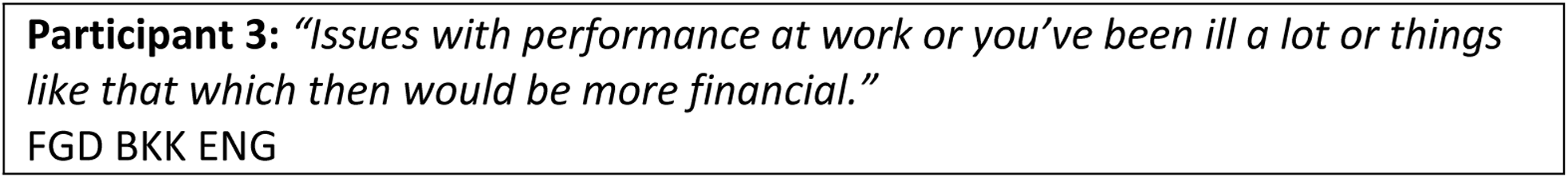

Social harms were also viewed as key indicators of problematic *hi-fun*. In Pattaya, men frequently described experiences of violence, coercion and being pressured into situations against their will, including interactions with police. Legal involvement, particularly arrest or mandated rehabilitation, was widely understood as a sign that *hi-fun* had escalated into a serious problem.

In addition, some participants recounted incidents of sexual violence, unwanted recording, or being forced to take unknown substances. These events were described as occurring when control was lost, either personally or within the group, and were considered clear markers that *hi-fun* had become unsafe.

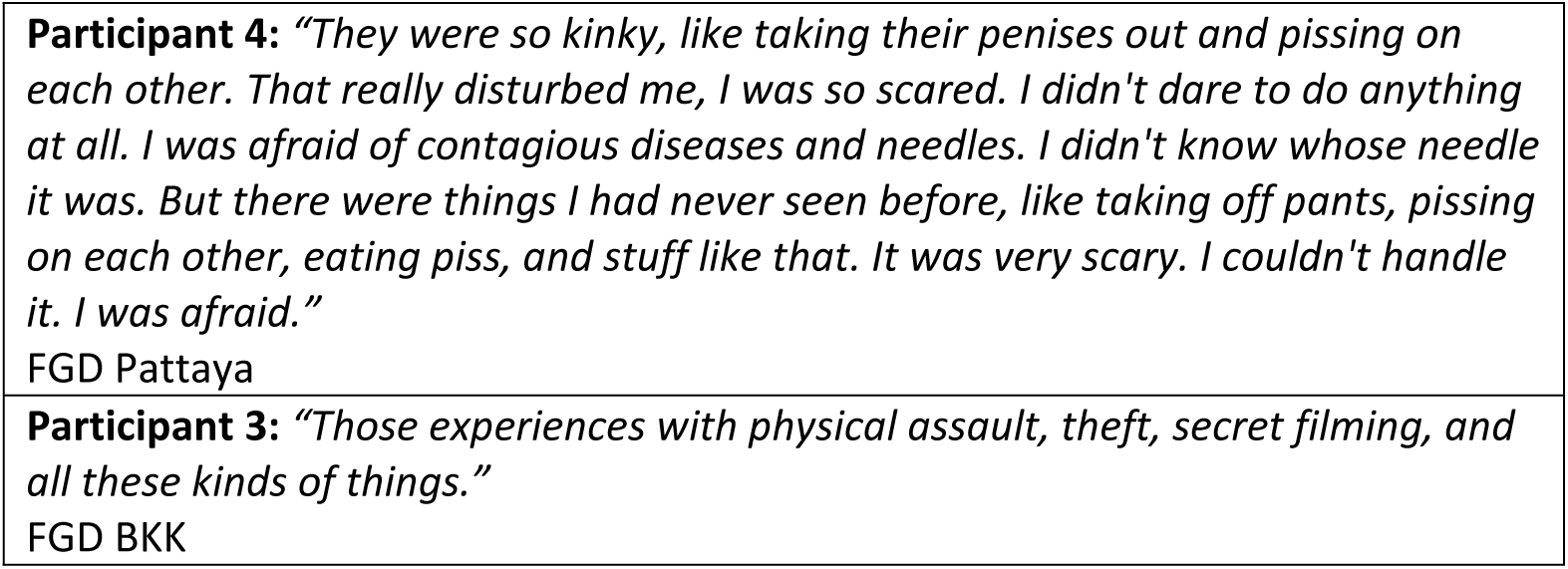

### Walking the line: Managing use and minimizing harm

Although participants recognized the risks associated with hi-fun, many described strategies to manage and reduce harm. Some viewed substances as bad but manageable when used with clear limits. Participants described avoiding certain drug combinations, such as poppers with ice, to reduce cardiac risks, and many set personal rules around frequency, often restricting *hi-fun* to once a month to prevent dependence, maintain pleasure and protect their health.

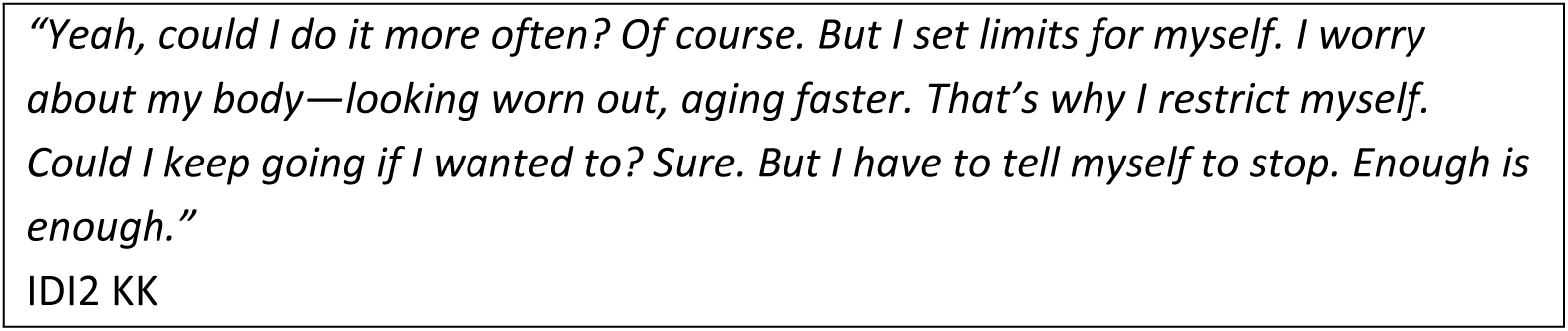

Participants also adopted measures to prevent overdose and manage emergencies. These included keeping contact information for harm reduction NGOs, informing trusted friends about session details and bringing their own needles to reduce risks of BBV and STI transmission. Such practices demonstrate how individuals actively mobilised available resources to protect themselves and others.

Legal risk management was another important concern. In Bangkok, some men carried NGO-issued identification or health documents to present during police stops. However, most emphasised avoiding possession of substances, especially at night when inspections were more common. Others described asking women friends to carry drugs because they were less likely to be searched. In Khon Kaen, strategies focused more on maintaining health, such as limiting sessions and monitoring the physical impacts of use. A few European participants believed they could pay bribes if confronted by police. These diverse strategies highlight that individuals were not passive within structural constraints but actively navigated them through personal and pragmatic adaptations.

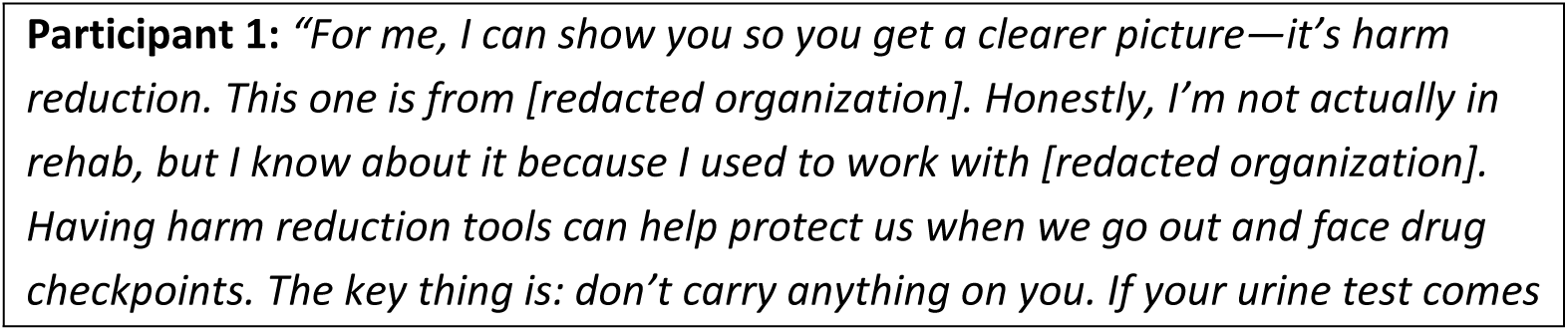
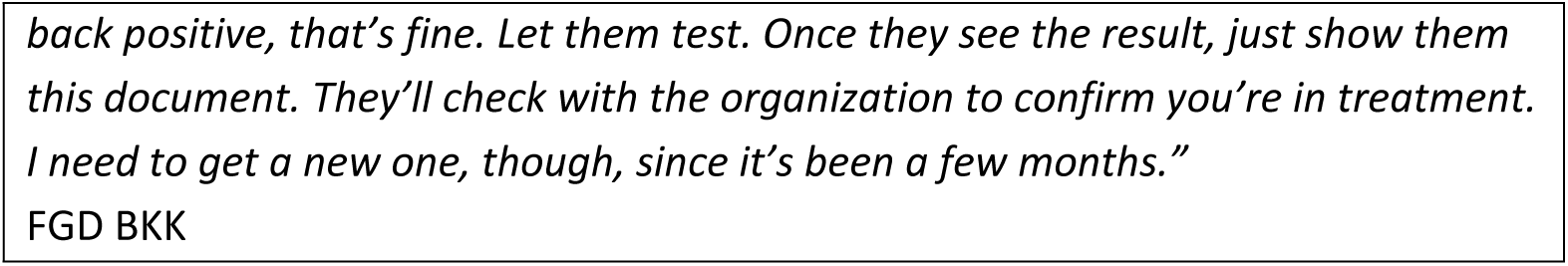

### What we need: Support beyond abstinence

The voices of participants emphasized that their needs extended beyond abstinence-based support. They highlighted the importance of resources that enable safer use and promote physical and emotional recovery. In Khon Kaen, men described needing vitamins, herbal detox remedies and post-use care, along with access to clean equipment and reliable information about safer practices. Some recounted having to reuse needles or clean them with water, saline or alcohol when supplies were unavailable, which they viewed as risky. These accounts reflect a harm reduction approach in which people seek tools and knowledge to care for their bodies in realistic conditions rather than through abstinence alone.

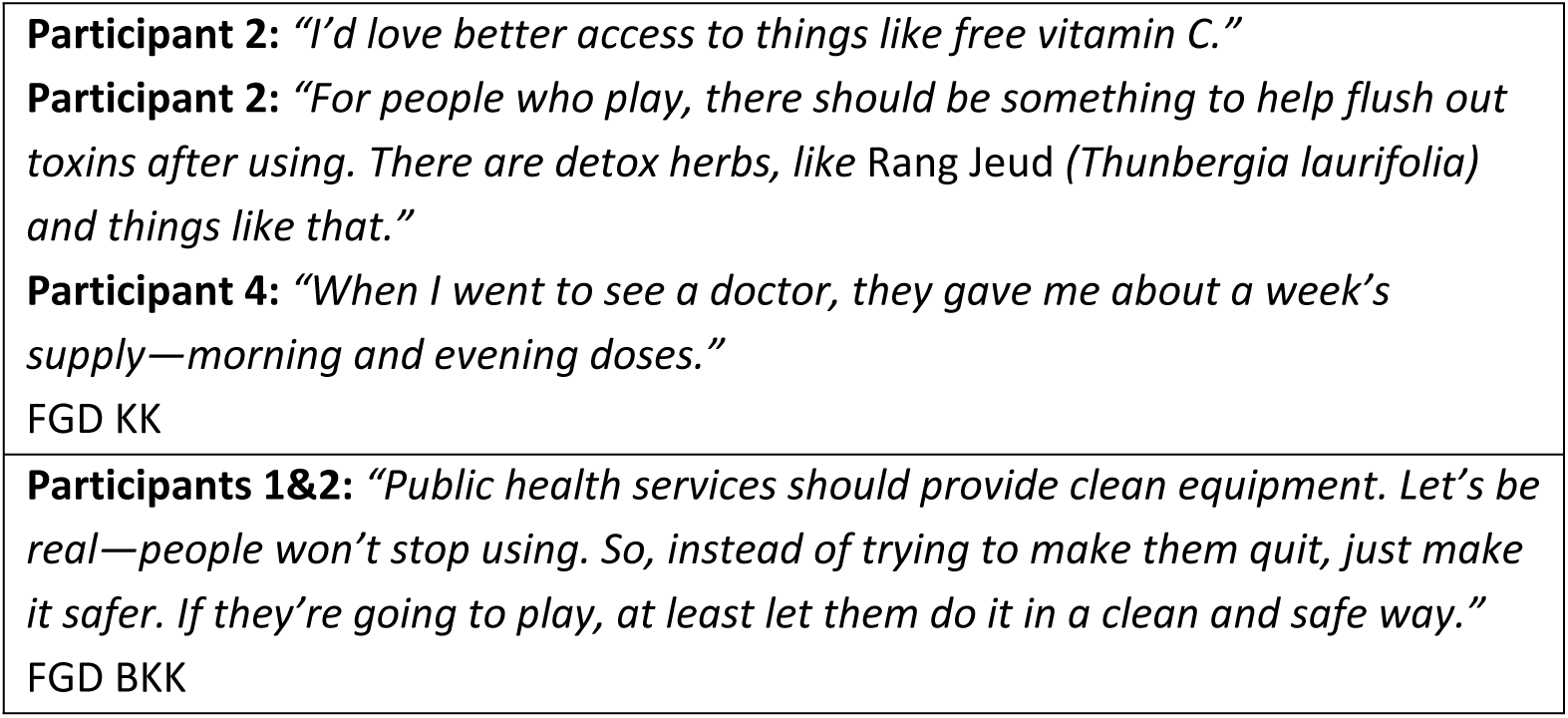

Participants also stressed the need for accurate and accessible information on safer drug use. They wanted practical guidance on mixing substances, safe injecting techniques and understanding drug interactions. While many actively sought to avoid harm, they often lacked non-judgmental and trustworthy sources of information. Existing abstinence-focused approaches were described as inadequate for addressing the realities of substance use.

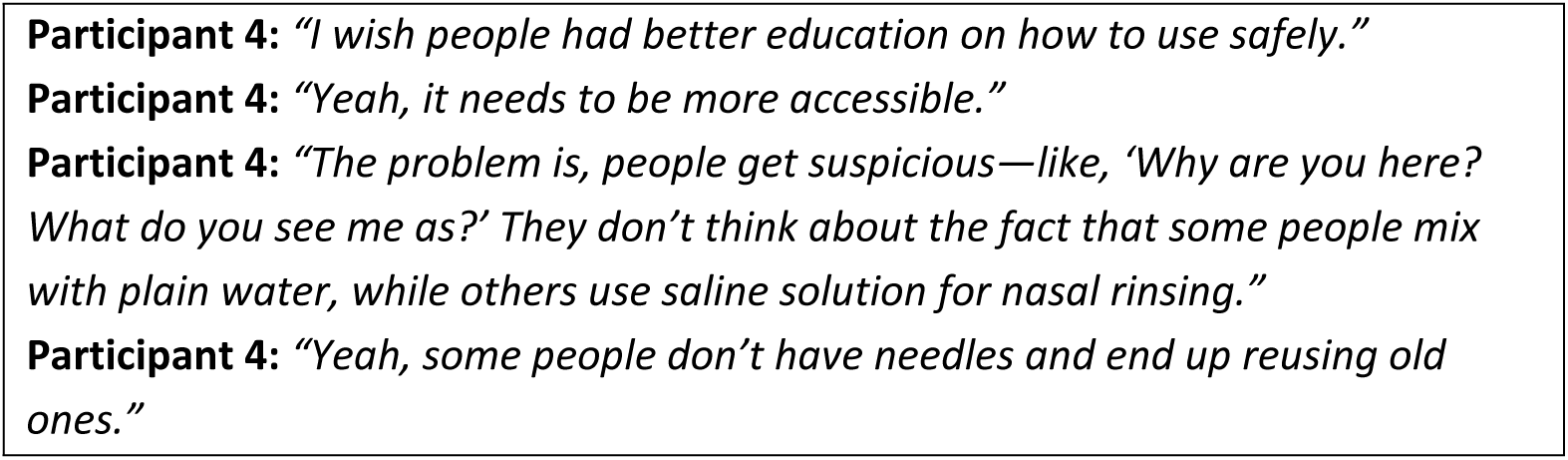
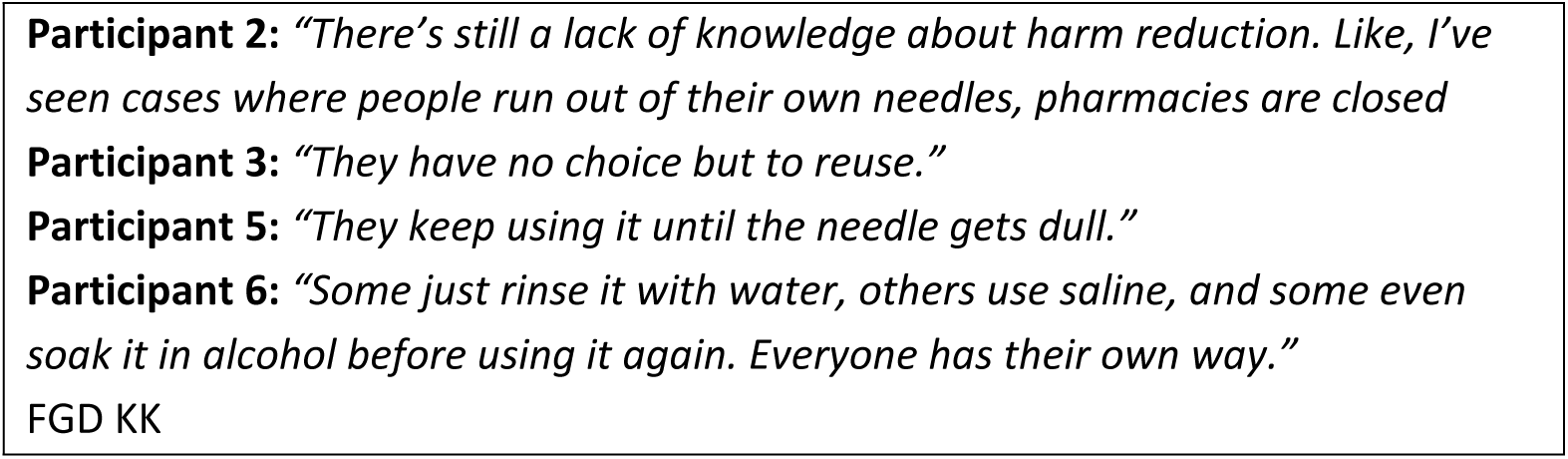

Beyond physical resources and knowledge, participants repeatedly highlighted the importance of acceptance and understanding from families, communities and society. Supportive, non-stigmatizing environments strengthened their ability to live well and manage use more safely.

Participants also expressed a need for broader structural change. They especially emphasized reform of legal and policing practices, noting that bribery, harassment and inconsistent enforcement created profound instability in their lives. Some men reported being fined or extorted even when they were not carrying substances but merely tested positive. They described legal thresholds for drug possession as unclear and misaligned with everyday patterns of use, contributing to fear and vulnerability.

These calls are not merely policy suggestions; they reflect a deeper demand for structurally safe spaces where individuals who use substances can care for themselves, access support, and live with dignity, without fear of prosecution, exploitation, or systemic abuse.

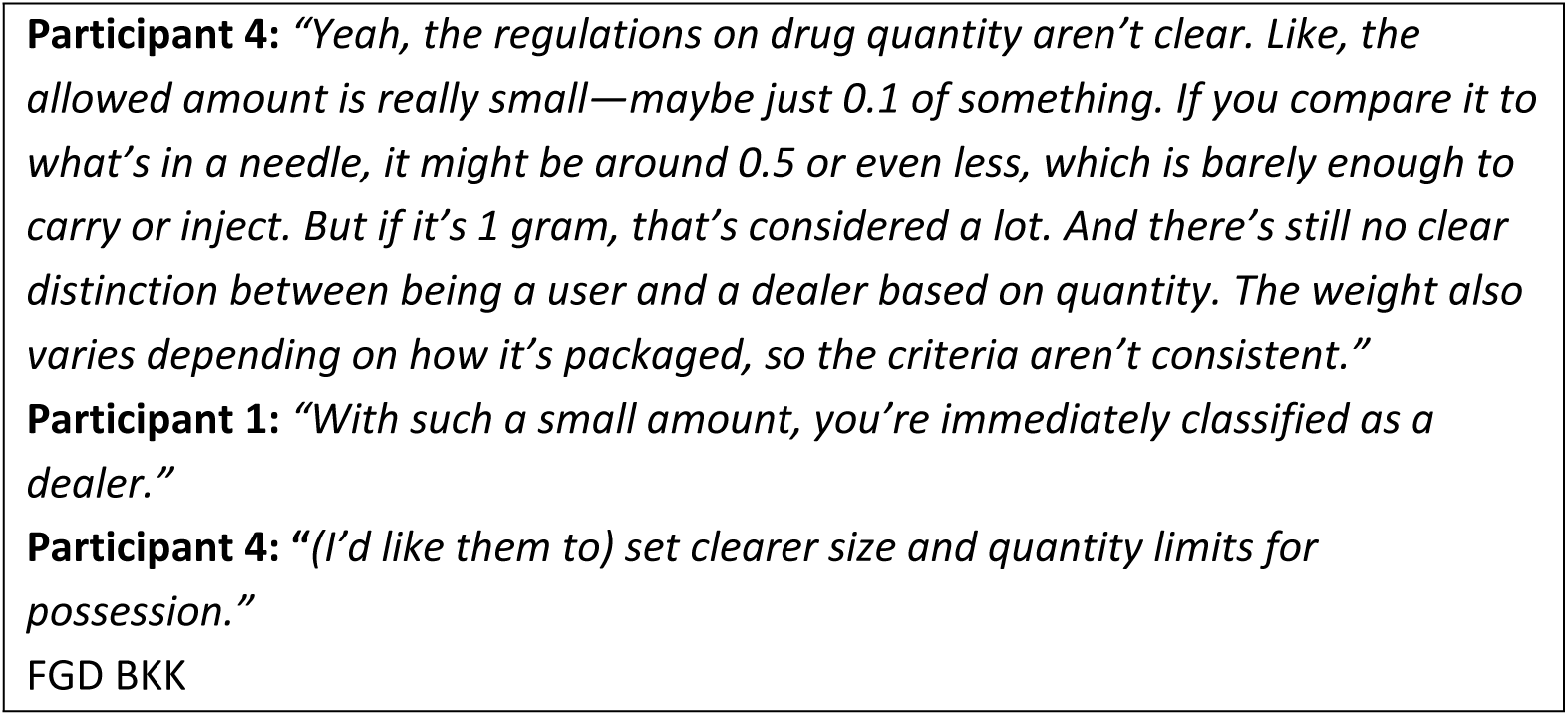

## Discussion

In this study, which included 30 GBMSM with experience of sexualised drug use, we found that the context of *hi-fun* was often described as a world of its own, where participants could feel free, be themselves, and express their sexuality and gender identity without restraint. *Hi-fun* was also portrayed as a carefully planned and organised setting, with strategies put in place to protect participants from potential harms. These strategies included carrying personal needles, screening individuals before inviting them to join to avoid police infiltration or theft and following shared rules such as prohibiting phone use during sessions except for inviting new participants or playing pornography. At the same time, *hi-fun* was viewed as a practice of release and relief, helping to reduce stress, ease loneliness, and heal emotional pain. From the perspective of participants, *hi-fun* was more than simply a risky behaviour as defined by the state (Vongchak et al., 2005). It was also a space of empowerment, emotional creation, healing, and identity formation, where they could experience acceptance in this imagined community. Yet this world was also interwoven with anxieties and risks, including dependency on substances, the perceived link between sex and drug use, difficulties with sexual performance, and the deterioration of both physical and mental health. These issues were framed as concerns but not necessarily problems so long as participants were able to maintain control.

Indeed, *hi-fun* became problematic precisely when control was lost. This included difficulties in regulating frequency of use, sustaining daily functioning and work performance, or facing external forces such as police, the law, and compulsory rehabilitation. Arrest and judicial processes were therefore described as clear markers that *hi-fun* had become a problem. Similarly, experiencing situations that were beyond their control, such as being subjected to violence, being secretly recorded, being coerced into unwanted sexual acts or excessive substance use, or being unable to recall what had happened during a session, were all cited as markers that *hi-fun* had become problematic. From these accounts, participants emphasised their desire for access to resources that would allow them to use substances more safely, to receive appropriate post-use care, and to gain knowledge relevant to their community. Most importantly, they expressed the need for legal reforms that recognise and respond to the realities of substance use, reflecting their call for a safer and more just environment in which their experiences could be understood and respected.

As demonstrated in this study of GBMSM’s experiences with *hi-fun* across three urban contexts in Thailand, *hi-fun* is not merely a site for drug-enhanced sexual activity, but a deliberately constructed world. It is designed to offer safety, privacy, and separation from mainstream societal scrutiny and normative moral frameworks. These spaces allow participants to express fluid identities, be themselves, and access forms of pleasure often denied in everyday life. This aligns with Bryant et al. (2018), who argue that sexual pleasure is a critical yet overlooked aspect within traditional health frameworks in Western chemsex research (Bryant et al., 2018). However, this study extends those understandings by showing how pleasure in Thailand’s *hi-fun* contexts is deeply intertwined with survival and the pursuit of legitimacy as citizens under perilous legal systems and while facing moral stigma. *Hi-fun* is not a boundlessly free space but one governed by unwritten rules and structured group discourses that form a distinct “scene” with its own logic (Drysdale, 2023; Race et al., 2023). This includes venue setup, preparation of injecting equipment, scheduled appointments, and the selection of suitable participants based on appearance and perceived bodily desirability.. These selection criteria reflect mainstream norms around beauty and masculinity are replicated in these seemingly countercultural settings.

Behavior in *hi-fun* is further regulated by informal rules aimed at fostering participation, such as bans on phone use during sessions or expectations that everyone would participate actively in sex and substance use. These norms powerfully shape experiences and define who is considered insider or outsider. This reflects Michel Foucault’s concept of heterotopia, a space outside normative life that reflects, resists, or generates experiences impossible elsewhere, yet remains materially and symbolically powerful. *Hi-fun* appears liberating but operates through subtle mechanisms of ordering, selection, and discursive control embedded in participants’ routines (Foucault, 2007; Foucault & Miskowiec, 1986). This is strongly supported by Nagington (2024), who challenges normative moral frameworks and describes chemsex as subcultures with their own moral orders and internal regulations. Power emerges not from external authorities but from mutual surveillance, adaptation, and collective consent among participants (Nagington, 2024).

Within these group-specific structures, *hi-fun* participants express diverse selves, not as pre-existing identities but as selves brought into being through performance and repetition (Butler, 2011). These performances include emotional engagement in activities, proactive roles for receptive partners (e.g., stimulating partners’ genitals), and creating a sexy, fun atmosphere aligned with *hi-fun* community norms. Through repetition, these selves become affirmed and desired within the space. Many participants report feelings of self-worth, healing, empowerment, being chosen, and emotional and sexual fulfillment experiences rarely accessible in daily life. *Hi-fun* thus emerges not only as a space of sexual pleasure but of identity construction, connection, and escape from external distress (Møller & Hakim, 2023).

Regarding risk and adversity, participants are not passive victims or powerless marginal subjects. Rather, they actively negotiate, manage risks, practice self-care, and strategically mobilize social, cultural, and bodily capital in response to structural constraints. These include unclear laws, moral stigma, and exclusion even within gay communities, where people who use drugs are often othered and cast outside the respectable mainstream. They are further constrained by biomedical frameworks that categorize them as “patients” or “deviants” (Florêncio, 2023; Mowlabocus, 2023).

Participants are thus not merely shaped by structure but act as agents who contest it. They create space, generate meaning, and sustain norms through practice, decisions, and relationship networks. These actions go beyond escaping structural violence (Farmer, 2004); they are claims to visibility and citizenship. This directly challenges biomedical discourses that reduce drug users to health problems requiring control (Nagington, 2024).

Meanwhile, external structures like state policy, legal frameworks, and health services exact structural violence by either failing to recognize *hi-fun* communities or by actively persecuting them. Dominant moral discourses draw lines between us and them, distinguishing those deemed worthy of support from those cast as criminal or needing rehabilitation. Biomedical definitions of drug users as patients or addicts not only categorize but serve to allocate power and distribute resources. Recognizing *hi-fun* participants as equal citizens would necessitate dismantling such frameworks and redistributing state resources, a challenge to entrenched systems that prioritize other populations. As a result, the state maintains its authority by rendering *hi-fun* invisible or illegitimate. Non-recognition, neglect, and resource denial function as a politics of control upheld by structural discourses, effectively erasing *hi-fun* participants and reinforcing inequitable distribution (Farmer, 2004; Foucault, 1978).

Therefore, supporting men engaged in *hi-fun* in Thailand must go beyond listening to their voices (Palmer et al., 2024; Tan et al., 2018; Tan, Phua, et al., 2021). It requires critically examining state resource allocation and affirming that *hi-fun* participants are citizens with dignity and human rights.

Findings from this study suggest that abstinence-based, compulsory treatment, and punitive legal-health models fail to meet the lived realities and well-being needs of *hi-fun* communities while also enacting structural violence. Participants clearly expressed the need for access to post-use body recovery resources (e.g., vitamins, detox herbs), clean equipment, and stigma-free spaces.

Drawing on this perspective, the findings of this study can be interpreted through the lens of critical medical anthropology, which understands health and illness as shaped, defined, and governed by political, economic, and social structures that contour the lived realities and everyday experiences of GBMSM engaged in hi-fun in Thailand. The results also illustrate how state institutions, legal frameworks, and moral discourses operate as forms of structural violence, determining who is deemed deserving of protection, who becomes visible, and who is pushed to the margins of health resource access. (Farmer, 2010)

Although some harm reduction and recovery services do exist, they are largely concentrated within private sectors and remain financially accessible only to middle/upper-middle class Thais or affluent foreigners (PULSE clinic, 2025). The absence of comparable, state-supported programs reflects not simply a resource gap but a deeper moral and ideological boundary in which sexualized drug use is cast as deviant rather than recognized as a legitimate public health issue. This moral framing limits the state’s willingness to adopt and invest in evidence-informed harm reduction strategies, thereby reinforcing structural inequities in access to healthcare.

These highlight the urgent need for community-driven harm reduction approaches (Tan, O’Hara, et al., 2021). Such models must break free from biomedical paradigms and moral ideologies centered on diagnosis, control, and punishment, and reject Western-centric frameworks misaligned with Southeast Asian realities. Policy advancement should begin with listening to the voices of local GBMSM communities, enabling them to articulate their own needs and experiences. These should be grounded in local knowledge, cultural practices, and community beliefs that have been sustained across generations while also recognizing the impact of globalization on Asian queer communities. Policy development, therefore, should adopt a holistic approach and prioritize what is realistically implementable, rather than adhering to state-defined ideals or importing frameworks from Western or other contexts that reflect external realities. Building on these principles, designing culturally sensitive strategies must go hand in hand with structural reforms, including legal reform to eliminate laws that criminalize sexualized drug use.

Some potentially implementable strategies include interventions that seek to increase the ability of individuals to self-advocate in *hi-fun* settings, programmes that harness and further strengthen mutual support among GBMSM engaged in *hi-fun*, and initiatives that provide safer drug-use information and materials, including resources for safer use (e.g., new injecting equipment) and holistic recovery (e.g., vitamins). Importantly, because *hi-fun* gatherings often involve advance planning and preparation, they offer multiple opportunities to intentionally embed harm reduction measures into the process itself. However, broader efforts must continue to address stigma towards people who use drugs, which remains pervasive and represents a significant barrier to service development in Thailand. Addressing this issue requires multi-level support that is grounded in understanding and acceptance from those around them, particularly family members and the broader society. A non-judgmental, non-stigmatizing, and non-discriminatory environment was viewed as a critical factor in strengthening their ability to live well and manage their drug use more effectively. This form of recognition and acceptance represents a deep dimension of social support, one that directly influences a person’s sense of self-worth. It can reduce feelings of isolation and help people who use substances feel that they still have a safe, respected place in society. Especially in contexts where drug use is framed through the lens of criminality or illness, the presence of spaces where they are accepted as equal, valuable, and dignified human beings becomes fundamental to building an inclusive and health-promoting society that truly leaves no one behind.

## Strengths and limitations

A key strength of this study lies in amplifying the voices of GBMSM who are genuinely engaged in *hi-fun*. The study also captures the diversity of local settings by covering three distinct research sites across Thailand. Moreover, participants represented a broad range of backgrounds in terms of ethnicity, education, and age, which allowed for insights into both similarities and differences across groups.

Nevertheless, this study has several limitations. Participant recruitment was primarily conducted through local NGOs, due to concerns about legal risks (see Witzel et al., 2025). As a result, all participants were individuals who had access to health services provided by these NGOs. Consequently, the study lacks the perspectives of GBMSM who are unable to access such services, or who remain hidden from NGO support systems.

The use of FGDs focused largely on discussing normative understandings around *hi-fun* contexts, rather than exploring personal and in-depth experiences. This limited the richness of data related to legal issues, such as arrest, incarceration, or engagement with rehabilitation programs within focus groups. Participants may have felt uncomfortable sharing sensitive personal experiences in a group setting, which resulted in some surface-level reflections.

Finally, although the study intended to include both cisgender men and transgender men, all participants recruited were cisgender men. The absence of transgender men participants may indicate contextual differences in *hi-fun* participation between these groups, which remains an important gap for future research.

## Conclusions

This research highlights the inherent complexities that coexist within GBMSM in Thailand’s *hi-fun* spaces. *Hi-fun* is experienced as a sphere of freedom, healing, empowerment, and authentic self-expression and serves as a site for building networks and relationships that enhance participants’ sense of self-worth. However, *hi-fun* is also deeply connected with subcultural norms and broader economic and legal forces which shape and sometimes constrain these experiences.

Within this world of mixed pleasures, many participants voiced underlying concerns such as losing control over their substance use, which could affect their work and their mental health. They also described situations where diminished self-control left them vulnerable to violence, coercion, or threats. For participants, these experiences signaled that *hi-fun* was becoming problematic, not just a challenge or a risk. The possibility of arrest, prosecution, or compulsory treatment was seen as intensifying this further.

Ultimately, what GBMSM in the *hi-fun* context most need is not the legislative action or stigmatization they currently face, but recognition of their humanity, genuine understanding, and meaningful support. This includes listening to their voices, creating environments free from discrimination, and ensuring access to resources that enable safer substance use. Most importantly, it calls for reforms in laws and policies that reflect lived realities, acknowledge people who use substances as human beings with dignity, and provide opportunities for them to live stable, safe, and lives free from marginalization.

## Data Availability

The data underlying the findings of this study are presented within the manuscript. The underlying qualitative data consist of interview transcripts, which cannot be made publicly available due to ethical, legal, and confidentiality restrictions to protect participant privacy.

## Acknowledgements

We would like to express our sincere gratitude to all participants for their valuable contributions to this research. We also thank our colleagues from ACTTEAM, IHRI, and HON for their kind support in the recruitment process.

## Author contributions

Conceptualization: WW, TW, AB, AR, TG

Data curation: WW, TW

Formal analysis: WW, TW

Funding acquisition: TW

Investigation: WW, TW, NS, AB, AR, HP, NP, SS, VN, AB, TG

Supervision: AR, HP, GL, NP, SS, VN, AB, TG

Writing – original draft: WW

Writing – review and editing: TW, AR, HP, GL, NP, SS, VN, AB, TG, NS

## Funding

This work was supported by the Wellcome Trust [226345/Z/22/Z]. TG was funded by the U.S. National Institute of Mental Health (R01MH119015 and R34MH123337) and Mahidol University (Fundamental Fund: fiscal year 2025 by National Science Research and Innovation Fund (NSRF)).

## Declarations

Ethical review was sought from, and granted by, the research ethics committee at the Mahidol University, Faculty of Social Sciences and Humanities (COA 2024/030.1902) as well as University College London (COA 24583/001). All participants provided verbal recorded consent.

## Consent for publication

All authors reviewed and approved the final manuscript.

## Notes

### Competing Interest Statement

The authors have declared no competing interest.

### Funding Statement

This work was supported by the Wellcome Trust [Grant number: 226345/Z/22/Z]. Thomas E. Guadamuz received funding from the U.S. National Institute of Mental Health (R01MH119015 and R34MH123337) and Mahidol University (Fundamental Fund, Fiscal Year 2025, supported by the National Science Research and Innovation Fund, NSRF). The funders had no role in study design, data collection and analysis, decision to publish, or preparation of the manuscript.

